# Differential effects of socio-demographic factors on maternal haemoglobin concentration in three sub-Saharan African Countries

**DOI:** 10.1101/2020.05.06.20082941

**Authors:** Dickson A. Amugsi, Zacharie T. Dimbuene, Catherine Kyobutungi

**Affiliations:** Maternal and Child Wellbeing Unit, African Population and Health Research Center, APHRC Campus, P.O Box 10787-00100, Nairobi, Kenya; Department of Population Sciences and Development, University of Kinshasa, Democratic Republic of the Congo; Social Analysis and Modeling Division, Statistics Canada, Ottawa, Canada K1A 0T6

**Keywords:** Haemoglobin concentration, socio-demographics, Africa, quantile regression

## Abstract

**Objective:** To investigate the effects of socio-demographic factors on maternal haemoglobin (Hb) at different points of the conditional distribution of Hb concentration.

**Methods:** We analysed the Demographic and Health Surveys data from Ghana, Democratic Republic of the Congo (DRC) and Mozambique, using Hb concentration of women aged 15-49 years as an outcome of interest. We utilise quantile regression to estimate the effects of the putative socio-demographic factors across specific points of the maternal Hb concentration.

**Results:** We observed crucial differences in the effects of socio-demographic factors along the conditional distribution of Hb concentration. In Ghana, years of schooling had a positive effect on Hb concentration of mothers in the 5^th^ and 10^th^ quantiles. A year increase in schooling was associated with a unit increase in Hb concentration across all quantiles in Mozambique, with the largest effect at the lowest quantile. In contrast, a year increase in maternal education was associated with a unit decrease in Hb concentration of mothers in the three upper quantiles in DRC. Maternal body mass index had a positive effect on Hb of mothers in the 5^th^, 10^th^, 50^th^ and 90^th^, and 5^th^ to 50^th^ quantiles in Ghana and Mozambique, respectively. Breastfeeding had a significant positive effect on maternal Hb concentration across all countries, with the largest effect occurring at the lower quantiles. All the household wealth indices had positive effects on Hb concentration across quantiles in Mozambique, with the largest effect among mothers in the upper quantiles. However, in Ghana, living in a poor wealth index was inversely related to Hb concentration of mothers in the 5^th^ and 10^th^ quantiles.

**Conclusions:** Our results showed that the effects of socio-demographic factors on maternal Hb concentration vary along its distribution. Interventions to address maternal anaemia should take these variations into account to identify the most vulnerable groups.

**What is already known?**

- Higher years of formal education consistently predict positive maternal Hb outcomes in OLS and logistic regression analysis.
- Low maternal BMI, parity, living in poorest wealth quintile and breastfeeding are inversely related to maternal Hb concentration.
- Evidence from logistic regression analysis suggests that mothers who live in better-off households tend to have a better Hb concentration.

**What are the new findings?**

- Socio-demographic factors have differential effects on Hb concentration at different points of its distribution.
- Interpreting results based on the mean effect (as in OLS) does not provide a comprehensive picture of the effects of the predictor variables.
- Breastfeeding has a positive effect on maternal Hb concentration contrary to the existing literature
- Contrary to the existing literature, maternal education is negatively associated with Hb of mothers in the three upper quantiles in DRC.

**What do the new findings imply?**

- Differential effects of socio-demographic factors on Hb concentration may help identify suitable interventions for groups most in need.
- The QR analysis suggests the need to look beyond OLS findings when designing interventions to improve maternal Hb concentration.
- Promoting breastfeeding among mothers may potentially improve their Hb concentration.
- The negative effect of maternal education on Hb in DRC, suggests that educational interventions to improve health outcomes should be context-specific

## Introduction

Maternal anaemia or low haemoglobin (Hb) concentration, a condition in which the Hb is lower than normal is a worldwide public health problem [1]. It is caused by deficiencies in iron, folate, copper, and other vital vitamins [2, 3]. Also, infectious disease morbidity, parasitic infections and blood related genetic disorders among others could cause low Hb concentration [3, 4]. While the causes of low Hb concentration are multifaceted, the evidence show that an estimated 50% of low Hb concentration cases reported worldwide are due to iron deficiency [5-7]. The available data suggest that anaemia affects about 500 million women of reproductive age, globally [8, 9]. The World Health Organisation (WHO) estimates indicate that the global anaemia prevalence >30% [2, 6]. Consequently, the WHO included a target of reducing anaemia among women of reproductive age by 50% by 2025 in its Global Nutrition Targets (GNT) [5]. Similarly, anaemia was recently added as an indicator to track the progress of sub-goal 2.2 of the Sustainable Development Goals (SDGs) to end all forms of malnutrition by 2030 [10]. It is significant to note that the problem of maternal low Hb concentration is particularly severe in sub-Saharan Africa (SSA) where poverty is highly prevalent and nutritious food is not easily accessible, coupled with high incidence of infectious diseases [11-13].

The consequences of low Hb concentration on the health of women include but are not limited to increased risks of low birth weight, preterm birth, perinatal mortality, and neonatal mortality [14]. Low Hb concentration also places women at an elevated risk of death during childbirth and postpartum [15]. Additionally, the literature suggests that deficient Hb concentration can result in cardiac decompensation (i.e. the failure of the heart to maintain adequate blood circulation). It also elevates the risk of haemorrhage and decreases the ability to tolerate blood loss, which can lead to circulatory shock and death [16, 17]. The consequences of low Hb concentration mentioned above calls for an investigation that would examine the effects of putative factors across the conditional distribution of the Hb concentration. Such an inquiry will provide entry points for interventions to address maternal anaemia in developing countries. This study intends to achieve this goal by using an analytical strategy that focuses on the effects of socio-demographic factors at different stages of the conditional distribution of maternal Hb concentration.

The existing literature has identified several factors that have both negative and positive effects on maternal Hb concentration. Some of these factors include, maternal age, education, parity, marital status, household size, socioeconomic status, place of residence, body mass index (BMI) and breastfeeding [18-25]. A study in Dhaka showed a strong relationship between maternal age, education level, income level, and maternal Hb concentration [18]. Moreover, higher BMI, primiparity, and living in better-off households were associated with higher levels of Hb [21, 22]. On the contrary, low family income and large family size are negatively associated with maternal Hb concentration [24]. Further, low maternal BMI, parity, living in poorest wealth quintile and breastfeeding were inversely related to maternal Hb concentration [25]. Other studies have shown that being separated or widowed, using an intrauterine device and place of residence increased the odds of low Hb concentration among women [22, 23].

Indeed, from the studies reviewed above, it appears the literature on maternal anaemia is abound. Nonetheless, there are shortfalls with the analytical strategies employed in these studies. For example, almost all these studies used either Ordinary Least Squares (OLS) or logistic regression to estimate the effects of socio-demographics on Hb concentration. This may mask the differential effects that may occur throughout the entire distribution of the Hb concentration--socio-demographic factors may influence maternal Hb differently across the distribution of the Hb concentration. The possible differences in the effects of socio-demographic factors on Hb concentration suggest the need to undertake an analysis that has the potential to present a comprehensive and complete picture of the effects of these factors on maternal Hb concentration. The quantile regression analytical strategy utilised in this paper can determine the effects of the socio-demographic factors at different points of the conditional distribution of maternal Hb concentration. Therefore, the objective this study was to examine the effects of sociodemographic factors on maternal Hb concentration using quantile regression. This type of analysis is currently missing in the anaemia research arena.

## Methods

### Data sources and study design

This study involved a secondary analysis of the demographic and health survey (DHS) data [26] from Ghana, Mozambique, and Democratic Republic of Congo (DRC). These are nationally representative data collected every five years in low and middle income countries (LMICs). We based the selection of the three countries on our previous analysis [27], as well as availability of anaemia data. In designing the surveys, the DHS ensured that the surveys are identical across all participating countries to facilitate comparison between and among nations. The DHS utilised a two-stage sample design in the selection of participating households in their surveys. The detail description of the DHS design processes is published elsewhere [28-32].

### Study participants

The study participants were women aged 15-49 years and who had complete anaemia data. Information on study participants was obtained through a face-to-face interviews. The DHS collected blood samples for anaemia testing from mothers who voluntarily consented to be tested [30]. Blood samples were drawn from a drop of blood taken from a finger prick and collected in a microcuvette. Hb concentration analysis was undertaken on-site using a battery-operated portable HemoCue analyser. Non-pregnant women with a Hb concentration less than 7.0 g/dl, and pregnant women with a Hb concentration lower than 9.0 g/dl were referred to a nearby health facility for immediate treatment [30]. The total samples per each country used in the present analysis were, Ghana (n= 2975), DRC (n= 9438) and Mozambique (n= 10961).

## Ethical statement

The DHS study was undertaken based on high ethical standards [33]. Data collectors were trained to recognise and respect the rights of study participants. They also informed participants of their rights to decide whether to participate in the study or not. The risks and benefits of the study as well as steps taken to mitigate the potential risks were adequately explained to study participants. The protocols of the study in each country, including biomarker collection, were approved by the recognised ethics review committees of each country, and the Institutional Review Board of ICF International, USA. Written informed consent was obtained from each study participant before they were allowed to participate in the study. The biomarker results were made available to study participants [30]. The DHS Program, USA, granted permission to the authors for the use of the data. Due to the anonymous nature of the data, the authors did not seek further ethical clearance.

## Measures

### Outcome variable

We used maternal Hb concentration (g/dl) as the outcome variable for this analysis. As described in the preceding sections, DHS collected blood samples from eligible women to test for anaemia using various strategies. The Hb concentration is captured first in the DHS data as a continuous variable and then categorised into three levels of anaemia: mild, moderate and severe. In this analysis, we used the Hb concentration as a continuous variable.

### Predictor variables

We grouped the predictor variables into three main categories: *maternal* (education, age, BMI, employment status, parity, breastfeeding status, marital status and ANC attendance); *household* (wealth index, sex of household head, household size, number of children under five years, decision making on large household purchases and husband/partner education) and *community* (place of residence). The DHS created the household wealth index using assets ownership and housing characteristics: type of roofing, and flooring material, source of drinking water, sanitation facilities, ownership of television, bicycle, motorcycle, automobile among others. The details of the computation processes is published elsewhere [30]. The maternal BMI (Kg/m^2^) was obtained by dividing weight in kilogrammes by height in meters squared.

## Data analysis

### Outline of quantile regression model

Koenker and Bassett [34] introduced the quantile regression (QR) as a *location model* to extend Ordinary Least Squares (OLS). It is the case because OLS summarises the distribution at the grand mean. However, the QR assesses more general class of linear models in which, the conditional quantiles have a linear form to fully account for the overall distribution of the response variable. To formalise the QR, we consider a real-valued random variable *Y* characterized by the following distribution function [34, 35];

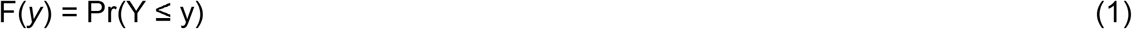

Then for any ☐ ☐ (0, 1), the ☐-th quantile of Y is defined as:

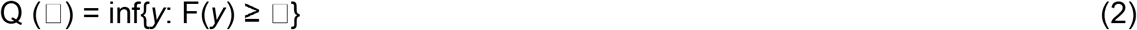

The common quantiles ☐ from Equation (1) are ☐ = .25, ☐ = .50, and ☐ = .75 for the first, quartile, the median and the third quartile. Therefore, unlike the OLS, which minimises the squared differences around the mean, QR minimises the weighted *absolute* difference between the observed value of y and the ☐-th quintile of Y. The preceding discussion demonstrates that OLS is nested within QR [34, 35].

## Analytical approach utilised

We used quantile regression (QR) [34] to examine the effects of the putative sociodemographic factors on maternal Hb concentration. Using the QR, we were able to investigate the effects of the predictor variables at different points of the conditional distribution of the outcome variable (Hb concentration). This type of analysis cannot be done with OLS, because standard OLS regression techniques summarise the average relationship between a set of regressors and the outcome variable based on the conditional mean function E (y|x). Thus, it provides only a partial view of the relationship, as we might be interested in describing the relationship at different points in the conditional distribution of y. The QR, unlike OLS, provides a complete view of the effects of the predictor variable on the outcome variable. Thus, making it possible to identify the vulnerable groups that are in dire need of interventions. Further, QR is more robust in handling non-normal errors and outliers compared with OLS [34]. Finally, QR provides a richer characterisation of the data, thereby illuminating the effects of a covariate on the entire distribution of the outcome variable. In this analysis, we also included OLS estimates for comparison purposes, and estimated QR at the 5^th^, 10^th^, 25^th^, 50^th^, 75^th^ and 90^th^ quantiles [27, 35].

Furthermore, since we did not have a specific predictor variable of interest, all the sociodemographic variables were included simultaneously in the models. They include, maternal education, age, BMI, employment status, parity, breastfeeding status, marital status, ANC attendance, household wealth index, sex of household head, household size, number of children under five years, decision making on large household purchases, husband/partner education and place of residence. The variables outlined above were selected based on the literature, followed by bivariate analysis. Significant variables in the bivariate analysis were included in the QR models.

## Results

### Descriptive analysis of the characteristics of samples

The descriptive results (Table 1) showed that mean maternal Hb concentration was relatively the same across all the three countries: Ghana (11.95±1.49), DRC (12.05±1.65) and Mozambique (11.64±1.73). Mothers in Ghana (5.27±4.92) spent a little more years in education than those in DRC (4.88±3.77) and Mozambique (3.46±3.45). The age of the study participants ranged from 28 years in Mozambique to 31 years in Ghana. Moreover, mothers in Ghana tended to have higher mean BMI (24.34±4.96) relative to those in DRC (21.79±3.66) and Mozambique (22.53±3.70). In DRC, 68% of mothers indicated they were breastfeeding at the time of the study, while the number of breastfeeding mothers in Ghana and Mozambique stood at 58%, respectively.

**Table 1:** Characteristics of the socio-demographic variables of the three countries

|  | Ghana (n=2975) |  | DRC (n=9438) |  | Mozambique (n=10961) |  |
| --- | --- | --- | --- | --- | --- | --- |
| Variables | Mean/% | SD | Mean/% | SD | Mean/% | SD |
| **Maternal-level variables** |  |  |  |  |  |  |
| Hb concentration (g/dl) | 11.95 | 1.49 | 12.05 | 1.65 | 11.64 | 1.73 |
| Women education (in years) | 5.27 | 4.92 | 4.88 | 3.77 | 3.46 | 3.45 |
| Age (in years) | 30.65 | 6.96 | 29.13 | 6.94 | 28.38 | 7.18 |
| Women's Body Mass Index (BMI)(Kg/m<sup>2</sup>) | 24.34 | 4.96 | 21.79 | 3.66 | 22.53 | 3.70 |
| Working (yes) | 79.3 |  | 76.0 |  | 37.5 |  |
| Parity | 3.62 | 2.17 | 4.42 | 2.56 | 3.8 | 2.28 |
| Breastfeeding (yes) | 57.9 |  | 67.8 |  | 57.6 |  |
| **Marital status** |  |  |  |  |  |  |
| Never married | 6.4 |  | 4.2 |  | 5.6 |  |
| Married or Cohabiting | 87.4 |  | 87.1 |  | 82.8 |  |
| Divorced/Widowed/Separated | 6.2 |  | 8.7 |  | 11.6 |  |
| **Household-level variables** |  |  |  |  |  |  |
| **Wealth index** |  |  |  |  |  |  |
| Poorest | 33.0 |  | 27.4 |  | 18.2 |  |
| Poor | 21.1 |  | 22.9 |  | 18.9 |  |
| Middle | 19.0 |  | 20.7 |  | 19.7 |  |
| Rich | 15.1 |  | 17.1 |  | 22.1 |  |
| Richest | 12.3 |  | 12.1 |  | 21.1 |  |
| Head of household (Female) | 23.7 |  | 22.2 |  | 32.9 |  |
| Household size | 5.80 | 2.82 | 6.74 | 2.86 | 6.16 | 2.79 |
| Number of children under five years | 1.73 | 0.93 | 2.16 | 1.04 | 1.86 | 0.98 |
| **Decision on household large purchases** |  |  |  |  |  |  |
| Respondent alone | 16.0 |  | 12.5 |  | 11.1 |  |
| Respondent and Husband/Partner | 46.5 |  | 50.3 |  | 57.1 |  |
| Husband/Partner alone | 24.1 |  | 36.7 |  | 30.7 |  |
| Someone else/Other | 13.4 |  | 0.59 |  | 1.1 |  |
| Husband education (in years) | 7.04 | 5.44 | 8.53 | 4.11 | 4.28 | 3.95 |
| **Community-level variables** |  |  |  |  |  |  |
| Place of Residence (% urban) | 39.8 |  | 28.7 |  | 32.1 |  |

SD: standard deviation; Hb: Haemoglobin; Note: SDs are reported only for continuous variables

### Quantile multivariable regression analysis of the effects of socio-demographic factors on maternal Hb concentration

In Tables 2-4, we present the QR results of the effects of socio-demographic factors on maternal Hb concentration in Ghana, DRC and Mozambique. We also reported the OLS estimates for the purposes of comparison with the QR results. The results from the OLS analysis showed that maternal education had strong positive effects on Hb concentration in Ghana and Mozambique. Thus, in both countries, one-year increase in education was associated with a unit increase maternal Hb concentration. A similar effects of BMI on Hb concentration were observed in all the three countries, so was breastfeeding practice.

However, in the QR analysis (Tables 2-4), the results revealed vital differences in effects at different points in the conditional distribution of maternal Hb concentration. For example, in Ghana, the effect of maternal years of education occurred at the first two lowest quantiles (5^th^ and 10^th^), with the largest effect at the 5^th^ quantile. Similarly, in Mozambique, one-year increase in maternal education was associated with a unit increase in Hb concentration across all quantiles, with the largest effect on mothers in the lowest quantile (5^th^) and the smallest effect at the highest quantile (90^th^). Interestingly, in DRC, maternal years of education had an inverse relationship with Hb concentration of mothers in the three upper quantiles. Thus, one-year increase in maternal years of education was associated with 0.015, 0.020 and 0.023 units decrease in maternal Hb concentration at the 50^th^, 75^th^ and 90^th^ quantiles, respectively. In Ghana, maternal BMI had a significant positive effect on Hb concentration of mothers in the 5^th^, 10^th^, 50^th^ and 90^th^ quantiles. The effect on the remaining two quantiles did not reach statistical significance. The positive effects of BMI on maternal Hb concentration was at the three lowest quantiles (5^th^, 10^th^ and 25^th^) in DRC, while in Mozambique, a unit increase in maternal BMI was associated with 0.031, 0.033, 0.029 and 0.018 units increase in maternal Hb concentration at the 5^th^, 10^th^, 25^th^ and 50^th^ quantiles, respectively. In each of the countries, the largest effect of BMI on maternal Hb concentration occurred at the lower end of the Hb distribution.

In Ghana, breastfeeding was positively and significantly associated with Hb concentration of mothers in the first four quantiles (5th, 10th, 25th, and 50th), with the least effect occurring at the 50th quantile. However, in DRC and Mozambique, breastfeeding had a decreasing effect across all quantiles in the respective countries. The largest effect in each country was at the lower end of the conditional distribution of the Hb concentration, while the smallest effect was at the higher end of the distribution. In Ghana, women participation in decision making regarding large household purchases was associated with a better Hb concentration among mothers in the 25th and 50th quantiles, while the partner taking the decision alone was associated positively with the Hb concentration at 5th, 25th and 50th quantiles. On the contrary, there was an inverse effect of the partner alone, deciding on large household purchases on Hb concentration of mothers in the 25th and 50th quantiles in DRC. The effects of female household headship in DRC was mixed. It associated positively with Hb concentration of mothers in the first three quantiles (5th, 10th and 25th), and negatively with the two upper quantiles (75th and 90th). In Mozambique, the household wealth index had a significant and increasing (from 5th to 90th) effect on maternal Hb concentration across almost all the quantiles. The largest effect occurred at the highest end of the Hb distribution (90th quantile). In Ghana, being in the lower wealth index was associated with a low Hb concentration among mothers in the 5th and 10th quantiles.318

**Table 2:**
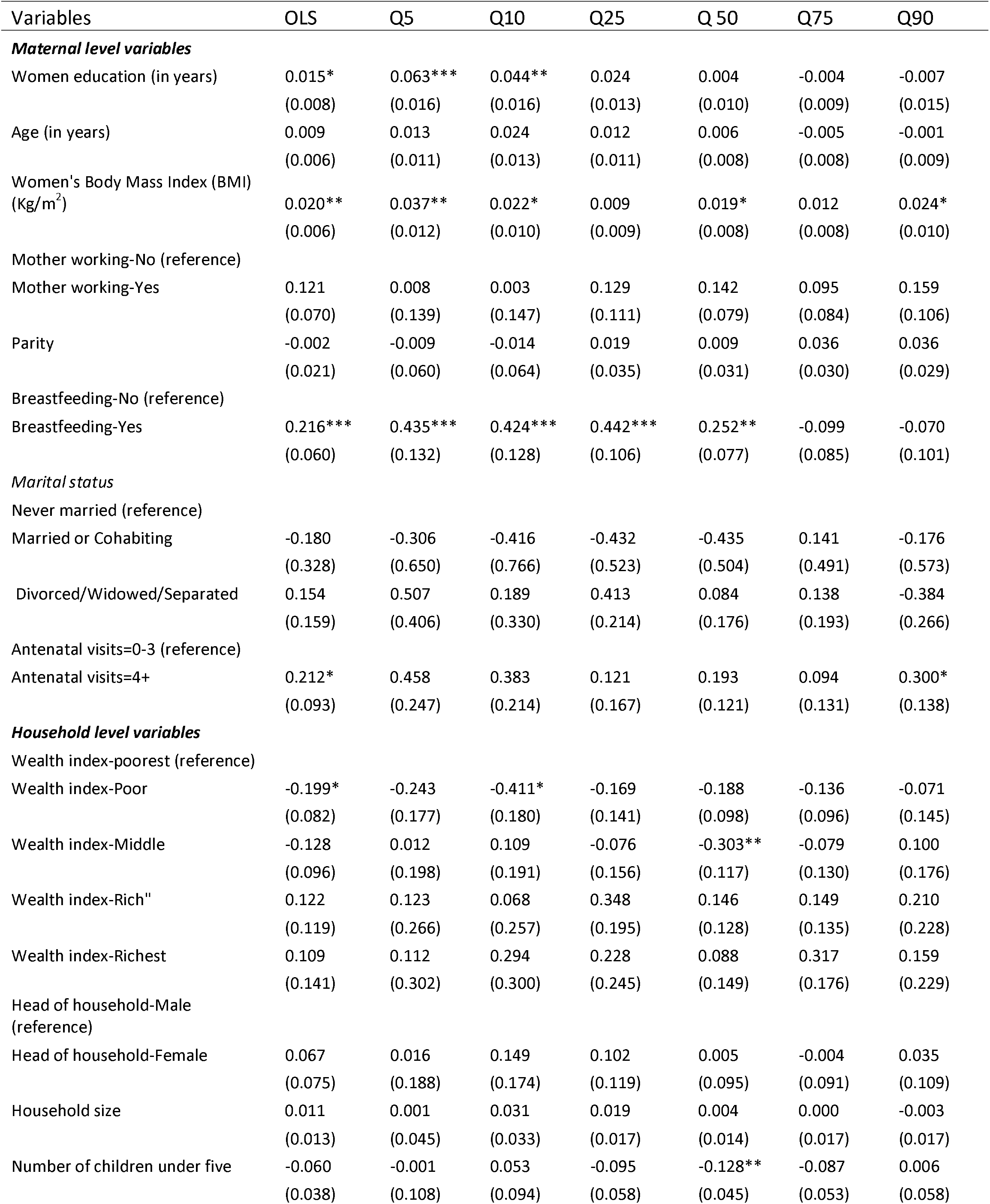
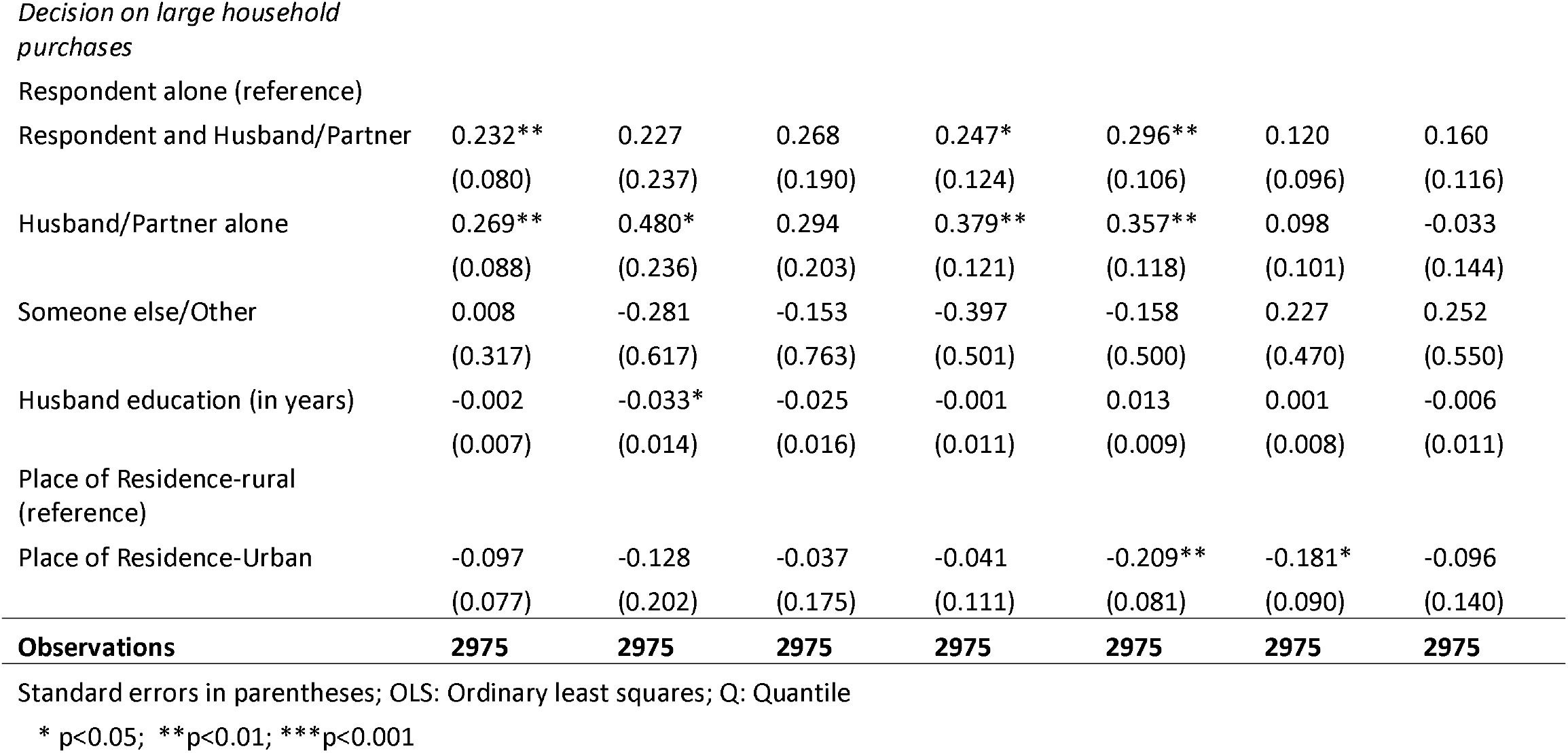
Effects of socio-demographic factors on maternal Hb concentration in Ghana

| Variables | OLS | Q5 | Q10 | Q25 | Q 50 | Q75 | Q90 |
| --- | --- | --- | --- | --- | --- | --- | --- |
| **Maternal level variables** |  |  |  |  |  |  |  |
| Women education (in years) | 0.015*<br>(0.008) | 0.063***<br>(0.016) | 0.044**<br>(0.016) | 0.024<br>(0.013) | 0.004<br>(0.010) | -0.004<br>(0.009) | -0.007<br>(0.015) |
| Age (in years) | 0.009<br>(0.006) | 0.013<br>(0.011) | 0.024<br>(0.013) | 0.012<br>(0.011) | 0.006<br>(0.008) | -0.005<br>(0.008) | -0.001<br>(0.009) |
| Women's Body Mass Index (BMI)<br>(Kg/m<sup>2</sup>) | 0.020**<br>(0.006) | 0.037**<br>(0.012) | 0.022*<br>(0.010) | 0.009<br>(0.009) | 0.019*<br>(0.008) | 0.012<br>(0.008) | 0.024*<br>(0.010) |
| Mother working-No (reference) |  |  |  |  |  |  |  |
| Mother working-Yes | 0.121<br>(0.070) | 0.008<br>(0.139) | 0.003<br>(0.147) | 0.129<br>(0.111) | 0.142<br>(0.079) | 0.095<br>(0.084) | 0.159<br>(0.106) |
| Parity | -0.002<br>(0.021) | -0.009<br>(0.060) | -0.014<br>(0.064) | 0.019<br>(0.035) | 0.009<br>(0.031) | 0.036<br>(0.030) | 0.036<br>(0.029) |
| Breastfeeding-No (reference) |  |  |  |  |  |  |  |
| Breastfeeding-Yes | 0.216***<br>(0.060) | 0.435***<br>(0.132) | 0.424***<br>(0.128) | 0.442***<br>(0.106) | 0.252**<br>(0.077) | -0.099<br>(0.085) | -0.070<br>(0.101) |
| **Marital status** |  |  |  |  |  |  |  |
| Never married (reference) |  |  |  |  |  |  |  |
| Married or Cohabiting | -0.180<br>(0.328) | -0.306<br>(0.650) | -0.416<br>(0.766) | -0.432<br>(0.523) | -0.435<br>(0.504) | 0.141<br>(0.491) | -0.176<br>(0.573) |
| Divorced/Widowed/Separated | 0.154<br>(0.159) | 0.507<br>(0.406) | 0.189<br>(0.330) | 0.413<br>(0.214) | 0.084<br>(0.176) | 0.138<br>(0.193) | -0.384<br>(0.266) |
| Antenatal visits=0-3 (reference) |  |  |  |  |  |  |  |
| Antenatal visits=4+ | 0.212*<br>(0.093) | 0.458<br>(0.247) | 0.383<br>(0.214) | 0.121<br>(0.167) | 0.193<br>(0.121) | 0.094<br>(0.131) | 0.300*<br>(0.138) |
| **Household level variables** |  |  |  |  |  |  |  |
| Wealth index-poorest (reference) |  |  |  |  |  |  |  |
| Wealth index-Poor | -0.199*<br>(0.082) | -0.243<br>(0.177) | -0.411*<br>(0.180) | -0.169<br>(0.141) | -0.188<br>(0.098) | -0.136<br>(0.096) | -0.071<br>(0.145) |
| Wealth index-Middle | -0.128<br>(0.096) | 0.012<br>(0.198) | 0.109<br>(0.191) | -0.076<br>(0.156) | -0.303**<br>(0.117) | -0.079<br>(0.130) | 0.100<br>(0.176) |
| Wealth index-Rich" | 0.122<br>(0.119) | 0.123<br>(0.266) | 0.068<br>(0.257) | 0.348<br>(0.195) | 0.146<br>(0.128) | 0.149<br>(0.135) | 0.210<br>(0.228) |
| Wealth index-Richest | 0.109<br>(0.141) | 0.112<br>(0.302) | 0.294<br>(0.300) | 0.228<br>(0.245) | 0.088<br>(0.149) | 0.317<br>(0.176) | 0.159<br>(0.229) |
| Head of household-Male<br>(reference) |  |  |  |  |  |  |  |
| Head of household-Female | 0.067<br>(0.075) | 0.016<br>(0.188) | 0.149<br>(0.174) | 0.102<br>(0.119) | 0.005<br>(0.095) | -0.004<br>(0.091) | 0.035<br>(0.109) |
| Household size | 0.011<br>(0.013) | 0.001<br>(0.045) | 0.031<br>(0.033) | 0.019<br>(0.017) | 0.004<br>(0.014) | 0.000<br>(0.017) | -0.003<br>(0.017) |
| Number of children under five | -0.060<br>(0.038) | -0.001<br>(0.108) | 0.053<br>(0.094) | -0.095<br>(0.058) | -0.128**<br>(0.045) | -0.087<br>(0.053) | 0.006<br>(0.058) |
| *Decision on large household purchases* |  |  |  |  |  |  |  |
| Respondent alone (reference) |  |  |  |  |  |  |  |
| Respondent and Husband/Partner | 0.232**<br>(0.080) | 0.227<br>(0.237) | 0.268<br>(0.190) | 0.247*<br>(0.124) | 0.296**<br>(0.106) | 0.120<br>(0.096) | 0.160<br>(0.116) |
| Husband/Partner alone | 0.269**<br>(0.088) | 0.480*<br>(0.236) | 0.294<br>(0.203) | 0.379**<br>(0.121) | 0.357**<br>(0.118) | 0.098<br>(0.101) | -0.033<br>(0.144) |
| Someone else/Other | 0.008<br>(0.317) | -0.281<br>(0.617) | -0.153<br>(0.763) | -0.397<br>(0.501) | -0.158<br>(0.500) | 0.227<br>(0.470) | 0.252<br>(0.550) |
| Husband education (in years) | -0.002<br>(0.007) | -0.033*<br>(0.014) | -0.025<br>(0.016) | -0.001<br>(0.011) | 0.013<br>(0.009) | 0.001<br>(0.008) | -0.006<br>(0.011) |
| Place of Residence-rural (reference) |  |  |  |  |  |  |  |
| Place of Residence-Urban | -0.097<br>(0.077) | -0.128<br>(0.202) | -0.037<br>(0.175) | -0.041<br>(0.111) | -0.209**<br>(0.081) | -0.181*<br>(0.090) | -0.096<br>(0.140) |
| **Observations** | **2975** | **2975** | **2975** | **2975** | **2975** | **2975** | **2975** |

Standard errors in parentheses; OLS: Ordinary least squares; Q: Quantile
\* p<0.05; \*\*p<0.01; \*\*\*p<0.001

**Table 3:** Effects of socio-demographic factors on maternal Hb concentration in DRC

| Variables | OLS | Q5 | Q10 | Q25 | Q 50 | Q75 | Q90 |
| --- | --- | --- | --- | --- | --- | --- | --- |
| **Maternal level variables** |  |  |  |  |  |  |  |
| Women education (in years) | -0.007<br>(0.006) | 0.016<br>(0.018) | -0.007<br>(0.013) | 0.006<br>(0.008) | -0.015**<br>(0.006) | -0.020**<br>(0.007) | -0.023*<br>(0.011) |
| Age (in years) | 0.004<br>(0.004) | -0.010<br>(0.012) | -0.007<br>(0.010) | 0.006<br>(0.005) | 0.012*<br>(0.005) | 0.005<br>(0.005) | 0.003<br>(0.006) |
| Women's Body Mass Index (BMI)(Kg/m<sup>2</sup>) | 0.015**<br>(0.005) | 0.026***<br>(0.008) | 0.021**<br>(0.007) | 0.014*<br>(0.006) | 0.007<br>(0.005) | 0.009<br>(0.007) | 0.001<br>(0.010) |
| Mother working-No (reference) |  |  |  |  |  |  |  |
| Mother Working -Yes | -0.033<br>(0.041) | 0.060<br>(0.107) | 0.046<br>(0.082) | -0.010<br>(0.052) | -0.037<br>(0.044) | 0.030<br>(0.051) | -0.038<br>(0.070) |
| Parity | 0.016<br>(0.012) | 0.052<br>(0.037) | 0.041<br>(0.026) | 0.017<br>(0.018) | 0.003<br>(0.014) | 0.014<br>(0.016) | 0.023<br>(0.018) |
| Mother breastfeeding-No (reference) |  |  |  |  |  |  |  |
| Mother breastfeeding-Yes | 0.470*** | 0.745*** | 0.662*** | 0.686*** | 0.488*** | 0.355*** | 0.219*** |
|  | (0.038) | (0.117) | (0.079) | (0.057) | (0.045) | (0.057) | (0.061) |
| *Marital status* |  |  |  |  |  |  |  |
| Never married (reference) |  |  |  |  |  |  |  |
| Married or Cohabiting" | -0.128 | -0.219 | -0.372* | -0.186 | -0.242** | -0.145 | 0.108 |
|  | (0.092) | (0.244) | (0.169) | (0.127) | (0.092) | (0.103) | (0.132) |
| Divorced/Widowed/Separated | 0.071 | -0.010 | -0.060 | -0.026 | 0.007 | 0.179 | 0.320* |
|  | (0.103) | (0.307) | (0.191) | (0.141) | (0.106) | (0.126) | (0.149) |
| Antenatal visits=0-3 (reference) |  |  |  |  |  |  |  |
| Antenatal visits=4+ | 0.025 | 0.011 | 0.027 | 0.030 | 0.014 | 0.016 | 0.095 |
|  | (0.039) | (0.098) | (0.078) | (0.053) | (0.043) | (0.045) | (0.063) |
| *Household level variables* |  |  |  |  |  |  |  |
| Wealth index-Poorest (reference) |  |  |  |  |  |  |  |
| Wealth index-Poor | -0.030 | 0.024 | 0.091 | -0.062 | -0.056 | -0.081 | -0.048 |
|  | (0.048) | (0.158) | (0.116) | (0.074) | (0.048) | (0.061) | (0.085) |
| Wealth index- Middle | 0.104* | 0.219 | 0.282* | 0.106 | 0.058 | -0.011 | 0.027 |
|  | (0.050) | (0.137) | (0.118) | (0.070) | (0.054) | (0.068) | (0.087) |
| Wealth index- Rich | 0.064 | 0.279 | 0.393** | 0.112 | -0.004 | -0.106 | -0.020 |
|  | (0.058) | (0.148) | (0.137) | (0.082) | (0.061) | (0.073) | (0.090) |
| Wealth index- Richest | -0.062 | 0.192 | 0.206 | -0.072 | -0.207* | -0.039 | -0.070 |
|  | (0.079) | (0.241) | (0.168) | (0.114) | (0.095) | (0.096) | (0.125) |
| Head of household-Male (reference) |  |  |  |  |  |  |  |
| Head of household-Female | 0.019 | 0.278* | 0.249** | 0.211*** | -0.033 | -0.129* | -0.179** |
|  | (0.045) | (0.136) | (0.083) | (0.057) | (0.046) | (0.057) | (0.068) |
| Household size | 0.001 | 0.035 | 0.001 | -0.004 | -0.002 | 0.002 | -0.009 |
|  | (0.008) | (0.020) | (0.016) | (0.012) | (0.011) | (0.010) | (0.011) |
| Number of children under five | -0.015 | -0.107 | -0.051 | 0.021 | -0.037 | -0.042 | 0.020 |
|  | (0.021) | (0.056) | (0.053) | (0.031) | (0.023) | (0.026) | (0.035) |
| *Decision on large household purchases* |  |  |  |  |  |  |  |
| Respondent alone (reference) |  |  |  |  |  |  |  |
| Respondent and Husband/Partner | -0.119* | -0.287 | -0.159 | 0.042 | -0.123 | -0.149* | -0.069 |
|  | (0.056) | (0.148) | (0.114) | (0.077) | (0.069) | (0.064) | (0.102) |
| Husband/Partner alone | -0.195*** | -0.235 | -0.243* | -0.103 | -0.250*** | -0.131 | -0.156 |
|  | (0.056) | (0.159) | (0.101) | (0.078) | (0.067) | (0.067) | (0.104) |
| Someone else/Other | 0.044 | 0.449 | 0.222 | -0.448 | 0.365 | -0.018 | -0.065 |
|  | (0.224) | (0.261) | (0.263) | (0.559) | (0.217) | (0.313) | (0.185) |
| Husband education (in years) | 0.005 | -0.005 | 0.002 | -0.010 | 0.006 | 0.011 | 0.019* |
|  | (0.005) | (0.015) | (0.012) | (0.007) | (0.005) | (0.006) | (0.009) |
| Place of Residence-Rural (reference) |  |  |  |  |  |  |  |
| Place of Residence-Urban | -0.004 | -0.124 | -0.132 | 0.027 | 0.131** | 0.018 | -0.140 |
|  | (0.051) | (0.138) | (0.108) | (0.069) | (0.049) | (0.056) | (0.093) |
| **Observations** | **9438** | **9438** | **9438** | **9438** | **9438** | **9438** | **9438** |

Standard errors in parentheses; OLS: Ordinary least squares; Q: Quantile
\* p<0.05; \*\*p<0.01; \*\*\*p<0.001

**Table 4:** Effects of socio-demographic factors on maternal Hb concentration in Mozambique

| Variables | OLS | Q5 | Q10 | Q25 | Q 50 | Q75 | Q90 |
| --- | --- | --- | --- | --- | --- | --- | --- |
| **Maternal level variables** |  |  |  |  |  |  |  |
| Women education (in years) | 0.035***<br>(0.007) | 0.055**<br>(0.021) | 0.043**<br>(0.015) | 0.030**<br>(0.011) | 0.037***<br>(0.008) | 0.046***<br>(0.009) | 0.027*<br>(0.011) |
| Age (in years) | 0.001<br>(0.004) | 0.002<br>(0.009) | 0.001<br>(0.007) | 0.001<br>(0.006) | 0.005<br>(0.004) | 0.000<br>(0.005) | 0.001<br>(0.005) |
| Women's Body Mass Index (BMI)(Kg/m<sup>2</sup>) | 0.019***<br>(0.005) | 0.031*<br>(0.015) | 0.033**<br>(0.011) | 0.029***<br>(0.007) | 0.018**<br>(0.006) | 0.008<br>(0.007) | 0.008<br>(0.008) |
| Mother working-No (reference) |  |  |  |  |  |  |  |
| Mother working-Yes | -0.049<br>(0.034) | 0.004<br>(0.094) | -0.038<br>(0.071) | -0.092<br>(0.052) | 0.001<br>(0.039) | -0.013<br>(0.042) | -0.080<br>(0.052) |
| Parity | 0.025*<br>(0.012) | 0.069*<br>(0.027) | 0.061**<br>(0.024) | 0.024<br>(0.015) | 0.003<br>(0.011) | 0.023<br>(0.017) | 0.004<br>(0.018) |
| Mother breastfeeding-No (reference) |  |  |  |  |  |  |  |
| Mother breastfeeding-Yes | 0.424***<br>(0.036) | 0.732***<br>(0.100) | 0.663***<br>(0.077) | 0.520***<br>(0.055) | 0.459***<br>(0.041) | 0.310***<br>(0.051) | 0.219***<br>(0.054) |
| **Marital status** |  |  |  |  |  |  |  |
| Never married (reference) |  |  |  |  |  |  |  |
| Married or Cohabiting | -0.067<br>(0.087) | -0.445*<br>(0.210) | -0.275<br>(0.160) | -0.159<br>(0.136) | 0.139<br>(0.106) | -0.055<br>(0.121) | 0.075<br>(0.134) |
| Divorced/Widowed/Separated | -0.139<br>(0.092) | -0.495*<br>(0.220) | -0.221<br>(0.205) | -0.144<br>(0.149) | -0.062<br>(0.113) | -0.138<br>(0.121) | -0.005<br>(0.141) |
| Antenatal visits=0-3 (reference) |  |  |  |  |  |  |  |
| Antenatal visits = 4+ | 0.049<br>(0.036) | 0.045<br>(0.086) | 0.028<br>(0.067) | 0.035<br>(0.048) | 0.015<br>(0.038) | 0.036<br>(0.041) | 0.075<br>(0.051) |
| **Household level variables** |  |  |  |  |  |  |  |
| Wealth index-Poorest (reference) |  |  |  |  |  |  |  |
| Wealth index-Poor | 0.255*** | -0.114 | 0.198 | 0.174* | 0.285*** | 0.326*** | 0.429*** |
|  | (0.054) | (0.157) | (0.110) | (0.076) | (0.063) | (0.063) | (0.074) |
| Wealth index- Middle | 0.367*** | 0.246 | 0.397*** | 0.249*** | 0.335*** | 0.399*** | 0.552*** |
|  | (0.054) | (0.131) | (0.109) | (0.073) | (0.055) | (0.067) | (0.075) |
| Wealth index-Rich | 0.380*** | 0.242 | 0.406*** | 0.268*** | 0.337*** | 0.400*** | 0.594*** |
|  | (0.056) | (0.146) | (0.118) | (0.079) | (0.059) | (0.067) | (0.085) |
| Wealth index-Richest | 0.257*** | -0.122 | 0.036 | 0.147 | 0.198** | 0.374*** | 0.474*** |
|  | (0.073) | (0.191) | (0.150) | (0.109) | (0.074) | (0.098) | (0.111) |
| Head of household-Male<br>(reference) |  |  |  |  |  |  |  |
| Head of household-Female | -0.010 | -0.061 | -0.150 | -0.040 | 0.075 | -0.050 | -0.074 |
|  | (0.039) | (0.100) | (0.083) | (0.057) | (0.045) | (0.047) | (0.067) |
| Household size | 0.015 | 0.014 | 0.007 | 0.004 | 0.028*** | 0.007 | 0.011 |
|  | (0.008) | (0.016) | (0.016) | (0.012) | (0.008) | (0.011) | (0.017) |
| Number of children under five | -0.025 | -0.077 | -0.070 | 0.001 | -0.053* | 0.015 | 0.003 |
|  | (0.022) | (0.054) | (0.051) | (0.035) | (0.024) | (0.034) | (0.034) |
| *Decision on large household purchases* |  |  |  |  |  |  |  |
| Respondent alone (reference) |  |  |  |  |  |  |  |
| Respondent and Husband/Partner | 0.010 | -0.146 | -0.092 | -0.050 | 0.075 | 0.027 | 0.088 |
|  | (0.056) | (0.143) | (0.106) | (0.084) | (0.064) | (0.062) | (0.093) |
| Husband/Partner alone | -0.032 | -0.021 | -0.061 | -0.031 | -0.050 | -0.020 | 0.043 |
|  | (0.059) | (0.149) | (0.112) | (0.088) | (0.065) | (0.069) | (0.099) |
| Someone else/Other | 0.244 | 0.817 | 0.349 | 0.133 | 0.062 | 0.430* | 0.314 |
|  | (0.168) | (0.605) | (0.230) | (0.267) | (0.227) | (0.172) | (0.260) |
| Husband education (in years) | 0.011* | -0.002 | 0.019 | 0.015 | 0.013 | 0.002 | 0.011 |
|  | (0.006) | (0.015) | (0.012) | (0.008) | (0.006) | (0.007) | (0.010) |
| Place of Residence-Rural<br>(reference) |  |  |  |  |  |  |  |
| Place of Residence-Urban | -0.059 | -0.278* | -0.203* | -0.184** | 0.019 | -0.021 | 0.139 |
|  | (0.046) | (0.129) | (0.088) | (0.067) | (0.046) | (0.058) | (0.072) |
| **Observations** | **10961** | **10961** | **10961** | **10961** | **10961** | **10961** | **10961** |

Standard errors in parentheses; OLS: Ordinary least squares; Q: Quantile
\* p<0.05; \*\*p<0.01; \*\*\*p<0.001

Figures 1-3 are visual presentations of the effects of the various socio-demographic factors on maternal Hb concentration in the three countries included in the analysis.

Figure 1: Pictorial presentation of the effects of socio-demographic factors on maternal Hb concentration in Ghana

Figure 2: Pictorial presentation of the effects of socio-demographic factors on maternal Hb concentration in DRC

Figure 3: Pictorial presentation of the effects of socio-demographic factors on maternal Hb concentration in Mozambique

## Discussion

This paper examined the effects of socio-demographic factors on maternal Hb concentration in Ghana, DRC and Mozambique, using quantile regression to understand the differential effects of putative socio-demographic factors at different points of the conditional distribution of the Hb concentration. Our QR results showed that in Ghana, one-year increase in maternal education was associated with an improved Hb concentration of mothers in the 5^th^ and 10^th^ quantiles, while the effects on the other four quantiles did not reach statistical significance. However, the OLS results show that one-year increase maternal education was associated with a unit increase in Hb concentration of all mothers. This paints just a part of the picture and therefore can be misleading. However, in Mozambique, the positive effect of years of schooling on Hb concentration was across all quantiles and in a decreasing manner. Thus, the largest effect of education occurred at the lowest quantiles, while the smallest effect was on Hb concentration of mothers in the highest quantile. Our findings in the two countries imply a disproportionate positive effects of maternal education accruing to mothers in the lower tail of the Hb distribution. This suggests that improving maternal education may be more impactful on the Hb concentration of mothers in the lower than the upper quantiles. Conversely, a year increase in maternal years of schooling was associated with a unit decrease in Hb concentration of mothers in the three upper quantiles in DRC. These findings are puzzling as the literature suggests that education consistently predict positive health outcomes in women [18, 25, 36, 37]. For example, a study using multicountry data concluded that women with higher years of education were less likely to be anaemic relative to those with fewer years of schooling [36]. Further research is needed to elucidate the possible factors accounting for the negative effect of maternal education on maternal Hb outcomes in DRC. Our study together with the above studies, despite using different analytical strategies, strongly suggest that education has positive effects on maternal health outcomes.

Our analysis also showed that maternal BMI has a significant positive effect on Hb concentration in at least three quantiles in each country. The most significant effect of BMI was among mothers in the lower quantiles in each country. This suggests that interventions targeted at improving maternal BMI qualitatively are likely to be more effective in increasing the Hb concentration of mothers in the lower tail of the Hb distribution. It is worthy to note that the effects of BMI were not across all quantiles. Hence, the OLS estimates, which suggested that a unit change in maternal BMI is associated with a unit increase in Hb concentration among all mothers, may be misleading. The QR findings, are, therefore, critical for identifying the groups that need to be targeted in programme planning. The literature corroborated the findings of our study. Several studies using either linear or logistic regression analytical strategies suggested that women with higher BMI tend to have higher levels of Hb concentration [22, 23, 36].

Similarly, breastfeeding had significant positive effect on maternal Hb concentration in all the three countries. Mothers who were breastfeeding at the time of the survey tended to have better Hb concentration compared with non-breastfeeding mothers. The largest effects were observed among mothers in the lower quantiles, suggesting that interventions to promote breastfeeding among lactating mothers may have more impact on Hb concentration of mothers at the lower end of the Hb distribution. These findings may appear puzzling because it is generally believed that lactating mothers tend to lose some iron to their infants, which may have a bearing on their Hb concentration [38, 39]. Nevertheless, other evidence suggests that the iron contained in breast milk to children is not significant enough to deplete the iron level of the mother, unless the mother is already anaemic [40]. The literature further suggests that mothers who are anaemic during pregnancy and postpartum can recover through a high intake of iron rich diet and/or iron supplement, and may not suffer low Hb concentration during lactation [40-42]. The preceding discussion suggests that breastfeeding may not necessarily deplete maternal iron level, with the consequential negative effect on Hb concentration. Some available evidence suggests a positive effect of breastfeeding on maternal Hb [38]. Nonetheless, other studies have observed inverse relationships between breastfeeding and Hb concentration levels [39, 43]. These mixed findings notwithstanding, the findings in the present study, together with the literature suggest that breastfeeding can indeed have positive effects on maternal Hb concentration levels, although the mechanism through which this happens may be complicated.

Our findings in Mozambique suggest that household wealth index (HWI) had a positive and increasing (from 5^th^ to 90^th^) effect on Hb concentration across all quantiles. The smallest effect was on mothers in the lower end of the Hb distribution, while the largest effect was on mothers at the upper end of the distribution. Thus, improving HWI may be more impactful on mothers at the upper quantiles relative to those at the lower quantiles. The positive association between HWI and women health outcomes have been substantially documented [38, 44, 45]. The evidence is that mothers who live in better-off households tend to have higher levels of Hb concentration [38]. However, in Ghana, mothers who live in poor households and are in the 5^th^ and 10^th^ quantiles tended to have lower Hb concentration. The finding in Ghana is consistent with the literature, which often identifies poverty as a risk factor of maternal health outcomes [25, 46].

An essential strength of this study is that the outcome variable was objectively measured, thereby reducing the possible biases associated with subjective measurements. The use of QR helped to examine the effects of the socio-demographic factors at different points of the Hb concentration, and thus present a comprehensive picture of the effects. Another necessary strength is the use of nationally representative data, making it possible for the results to be generalised to all women of reproductive age in the respective countries. We could not establish causality in this study due to the cross-sectional nature of the data. Also, missing data is an essential limitation of secondary data analysis. However, due to the robust measures put place by DHS to ensure the completeness of their datasets, missing data was not an issue in our study.

## Conclusions

We used quantile regression to examine the effects of socio-demographic factors on maternal Hb concentration. Our analysis demonstrated substantially that the various putative socio-demographic factors have differential effects on maternal Hb concentration at different points of the Hb distribution in all countries. Interventions and programmes to address maternal anaemia must take into account the different effects of the various socio-demographic factors on Hb concentration throughout the different percentiles of the Hb distribution. It may help identify suitable interventions for groups most in need.

## Data Availability

This study was a re-analysis of existing data that are publicly available from The DHS Program at http://dhsprogram.com/publications/publication-fr221-dhs-final-reports.cfm. Data are accessible free of charge upon registration with the Demographic and Health Survey program (The DHS Program). The registration is done on the DHS website indicated above.

## Acknowledgements

We wish to express our gratitude to The DHS Program, USA, for providing us access to the data. We also want to acknowledge the institutions of respective countries that played critical roles in the data collection processes.

## Authors’ Contribution

DAA conceived and designed the study, interpreted the results, wrote the first draft of the manuscript, and contributed to the revision of the manuscript. DAA and ZTD analysed the data. ZTD and CK contributed to study design, data interpretation, and critical revision of the manuscript. All authors take responsibility for any issues that might arise from the publication of this manuscript.

